# Early Detection of Diabetic Neuropathy Based on Health Belief Model: A Scoping Review

**DOI:** 10.1101/2023.12.15.23299437

**Authors:** Okti Sri Purwanti, Nursalam, Moses Glorino Rumambo Pandin

## Abstract

**Introduction:** Uncontrolled blood sugar levels may result in complications, namely diabetic neuropathy. Diabetic neuropathy is a nerve disorder that causes symptoms of numbness, foot deformity, dry skin, and thickening of the feet. The severity of diabetic neuropathy carries the risk of developing diabetic ulcers and amputation. Early detection of diabetic neuropathy can prevent the risk of diabetic ulcers.

**Methods:** This research studied articles in 6 databases via Scopus, Ebsco, Pubmed, Sage journal, Science Direct, and Springer link with the keywords “screening Neuropathy” AND “Detection Neuropathy” AND “Scoring Neuropathy” AND “Diabetic” published in 2019-2023. This study identified articles based on the English language, variables, and population. This research used PRISMA Flow-Chart 2020 to record the article filtering process.

**Results:** This research identified articles through PRISMA Flow-Chart 2020, obtaining 25 articles that discussed early detection of diabetic neuropathy.

**Conclusion:** Early detection of diabetic neuropathy prevents the occurrence of diabetic neuropathy. The method for early detection of neuropathy is using a neuropathy questionnaire and physical examination. Neuropathy questionnaires are Michigan Neuropathy Screening Instrument (MNSI), Toronto Clinical Neuropathy Score (TCNS), and Diabetic Neuropathy Classification (BDC). Physical examination tools include biotesimeters, vibrations, neurotesimeters, tuning forks, monofilament threads, ultrasonography (USG) results, artificial intelligence (AI), and nomograms.

## Introduction

Diabetes mellitus may cause complications of diabetic, retinopathic, and nephrotic neuropathy. Diabetic neuropathy is the most common complication in type 2 diabetes mellitus (Akhtar *et* al., 2023). Diabetic neuropathy occurs in the sensory, motor, and autonomic nervous systems (Saranya *et* al., 2019). Patients experience decreased sensitivity in the feet and legs. The severity of diabetic neuropathy causes diabetic wounds and ulcers (Lian *et* al., 2023). The health belief model identifies behaviors to prevent diabetes mellitus complications (Wang *et* al., 2023).

The health belief model considers perceived vulnerability, perceived benefits, perceived barriers, perceived seriousness, and received support (Orbell, 2020). Nursing services apply philosophy in providing nursing care. Philosophy has three main points of thought: ontology, epistemology, and axiology (Rokhmah, 2021).

The incidence of neuropathy in the world reaches 2.4% of the world’s human population, and the prevalence of neuropathy cases increases in old age by 8.0%. Globally, the highest prevalence of neuropathy occurs in the Asian continent. A higher incidence of neuropathy can be found in countries on the Southeast Asian continent, namely Malaysia (54.3%), the Philippines (58.0%) and Indonesia (58.0%) (Malik *et* al., 2020). A study shows that 50% of patients aged > 60 years’ experience neuropathy in the early stages of type 2 diabetes (Chevtchouk *et* al., 2017). In Indonesia, diabetes patients who experience complications from diabetic neuropathy reach 54% (Kementerian Kesehatan et al., 2020).

Early detection of neuropathy is to establish an early diagnosis and determine patient care. Proper treatment of neuropathy patients can prevent diabetic ulcers (Basir *et* al., 2020). Nurses can detect neuropathy early by using neuropathy instruments before signs of neuropathy symptoms appear. Patients aware of the signs of neuropathy can carry out appropriate foot care to prevent diabetic ulcers (Jones *et* al., 2023). Patients are willing to undergo a neuropathy examination if the patient feels the severity of neuropathy symptoms. Health workers diagnose neuropathy after clinical signs of neuropathy appear (Ziegler *et* al., 2022).

Based on the explanation above, early detection of neuropathy is carried out to confirm the diagnosis and prevent diabetic ulcers. This research aims to determine the early detection of diabetic neuropathy based on the health belief model.

## Methodology

This research used a systematic review research method. This research explained a review of ontology, epistemology, and axiology regarding early detection of diabetic neuropathy based on the health belief model. This research was conducted by collecting articles based on “screening Neuropathy,” “Detection Neuropathy,” “Scoring Neuropathy,” AND “Diabetic” through the databases PubMed, Scopus, Sage Journal, Science Direct, Ebsco, and SpringerLink. This research used articles in English and was published in the last five years (2019-2023). In this study, the process of selecting articles and recording used the PRISMA Flow-Chart 2020 diagram. In this panel, the article’s inclusion criteria were a population of diabetic neuropathy patients with early neuropathy detection interventions. The article selection process used rayyan.ai based on duplicate articles, abstracts, and types of research. The exclusion of articles was based on review articles, non-diabetes mellitus patient populations, book chapters, and articles appropriate to the topic, namely early detection of diabetic neuropathy.

## Results

This research conducted a journal search through 6 journal databases and filtered articles using PRISMA-P 2020. This research focused on articles discussing early detection of diabetic neuropathy. Based on filtering using PRISMA-P 2020, five articles were found. The article filtering process’s images were included in PRISMA-P 2020 below (figure 1).

**Figure 1.**
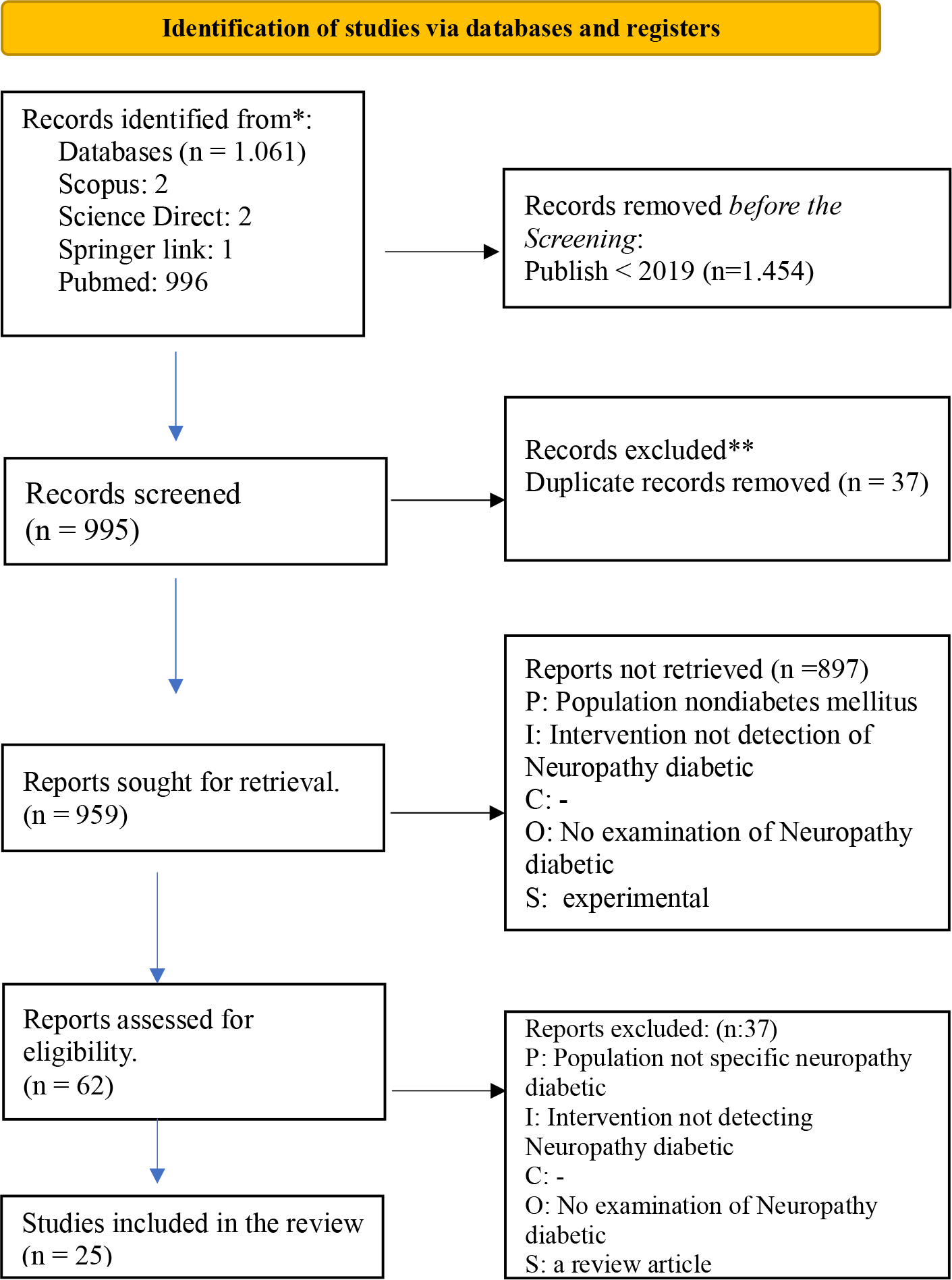
Flow chart Diagram **Source:** (Moher *et* al., 2015)

**Table 1.**
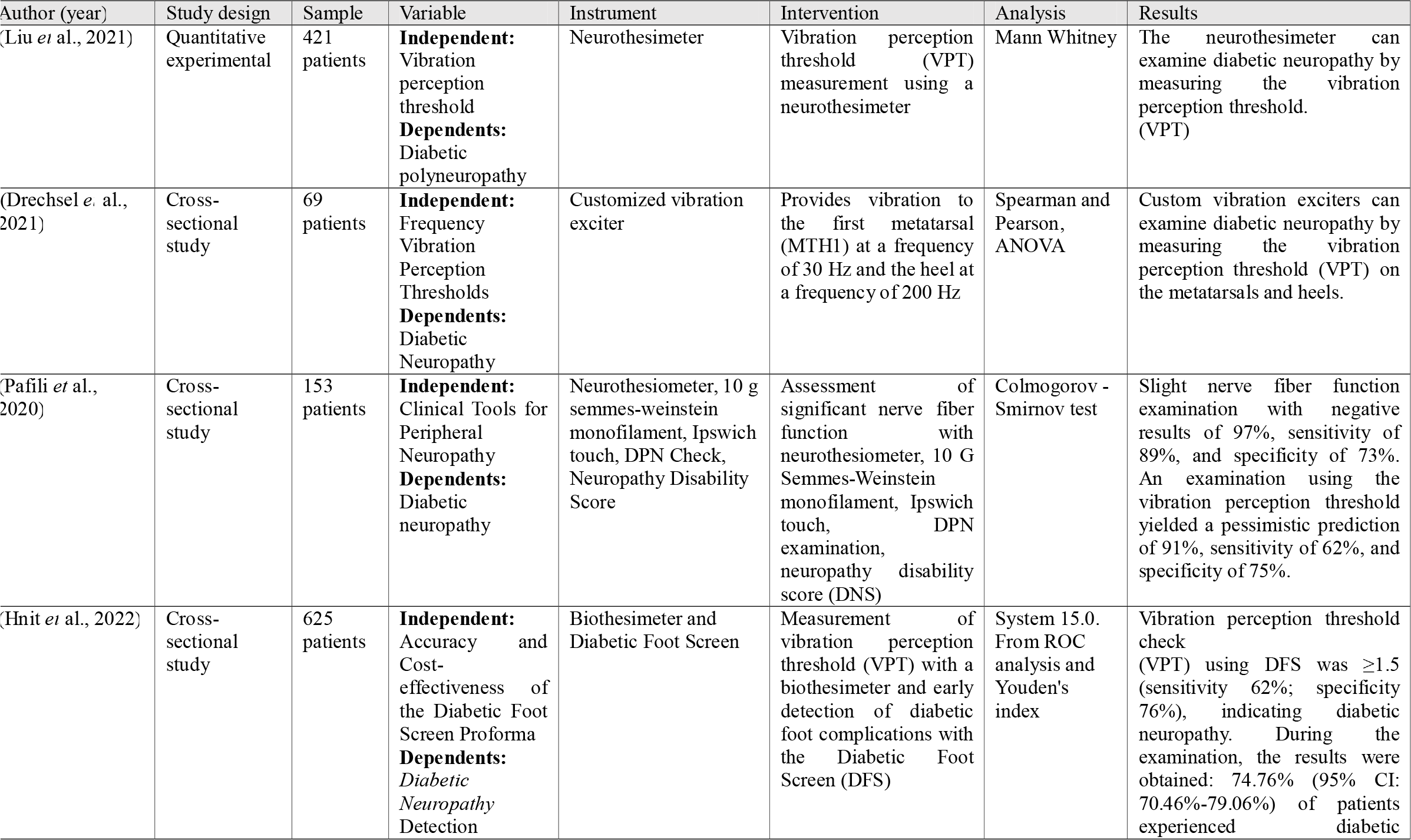
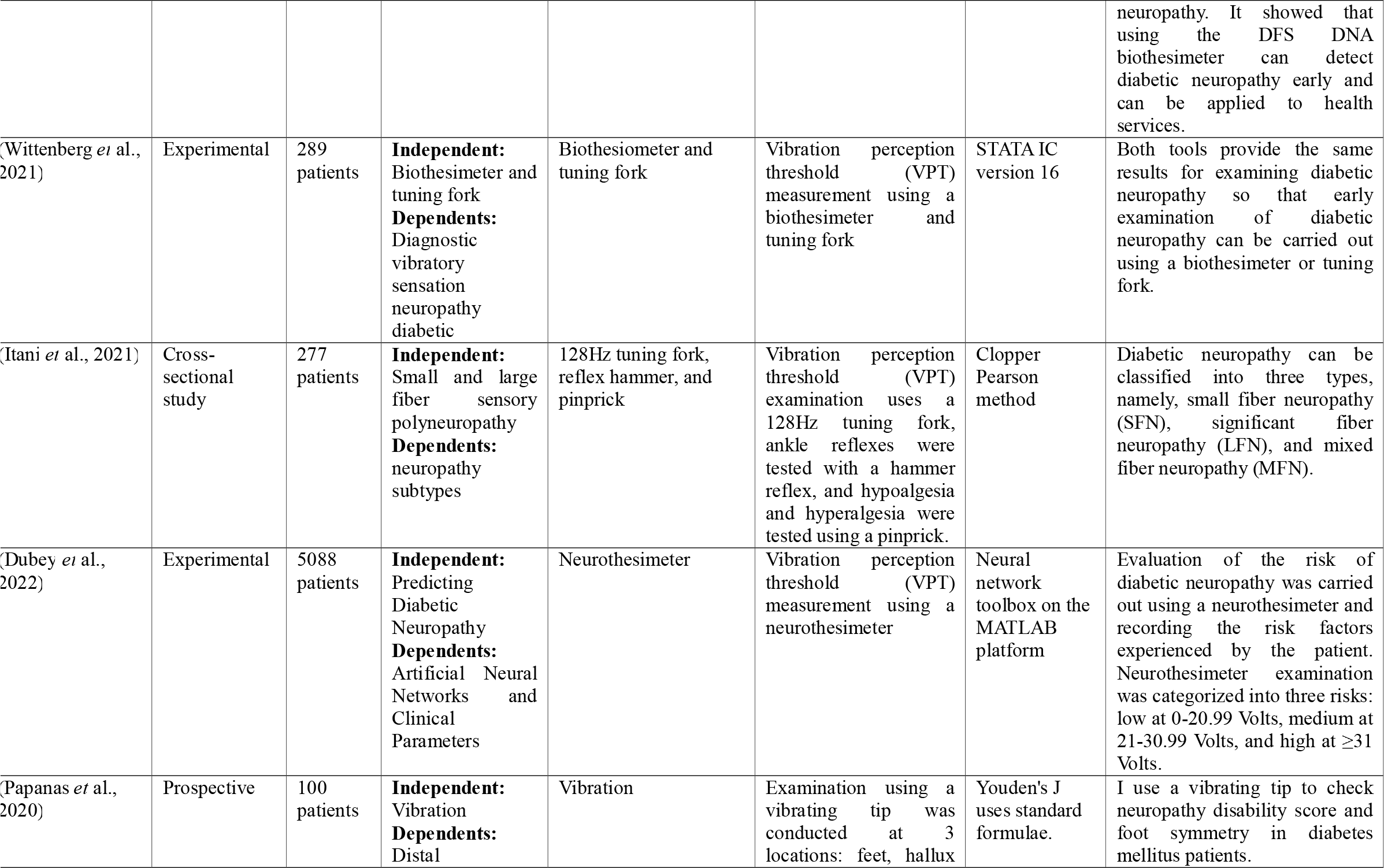
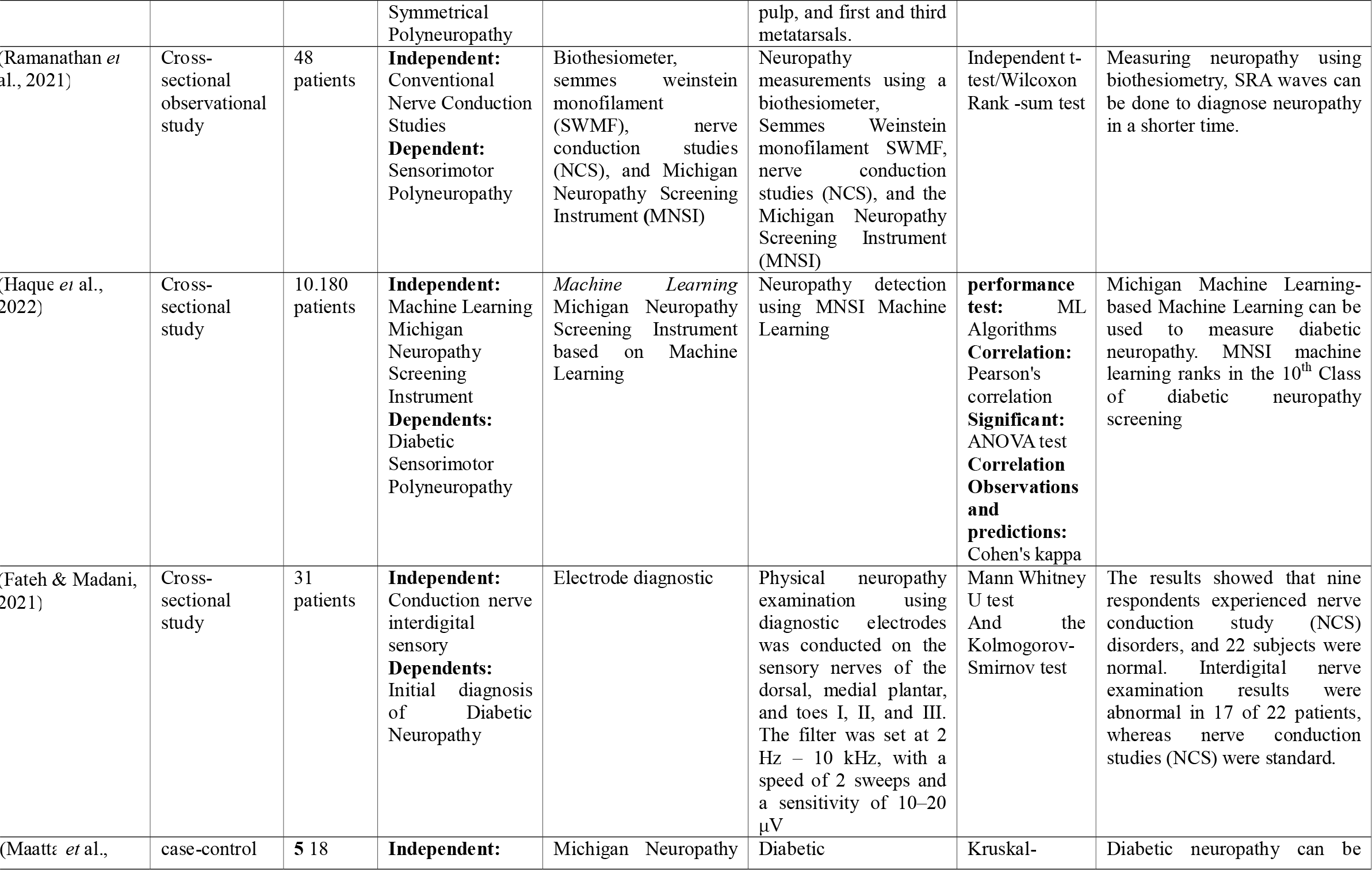
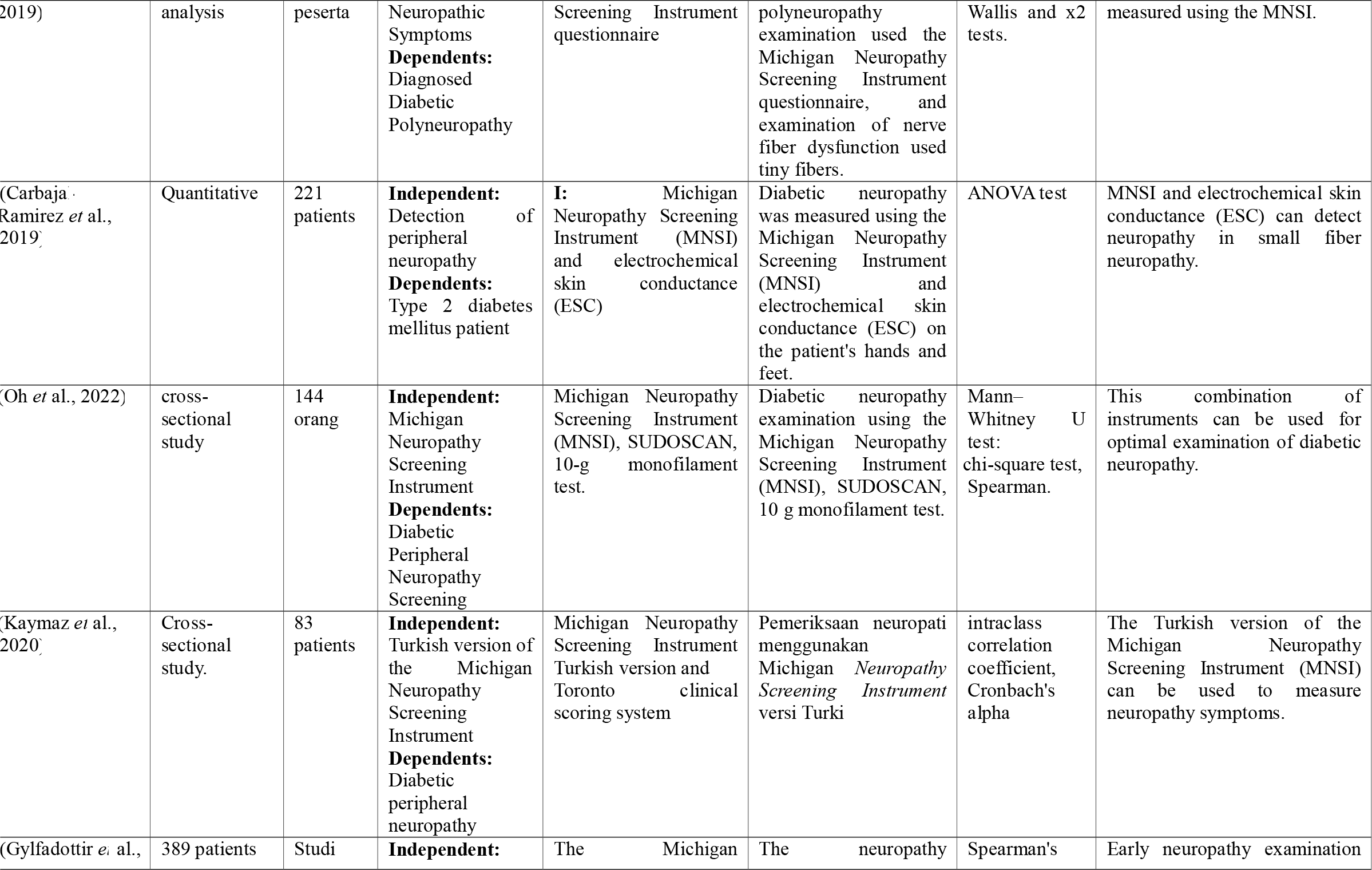
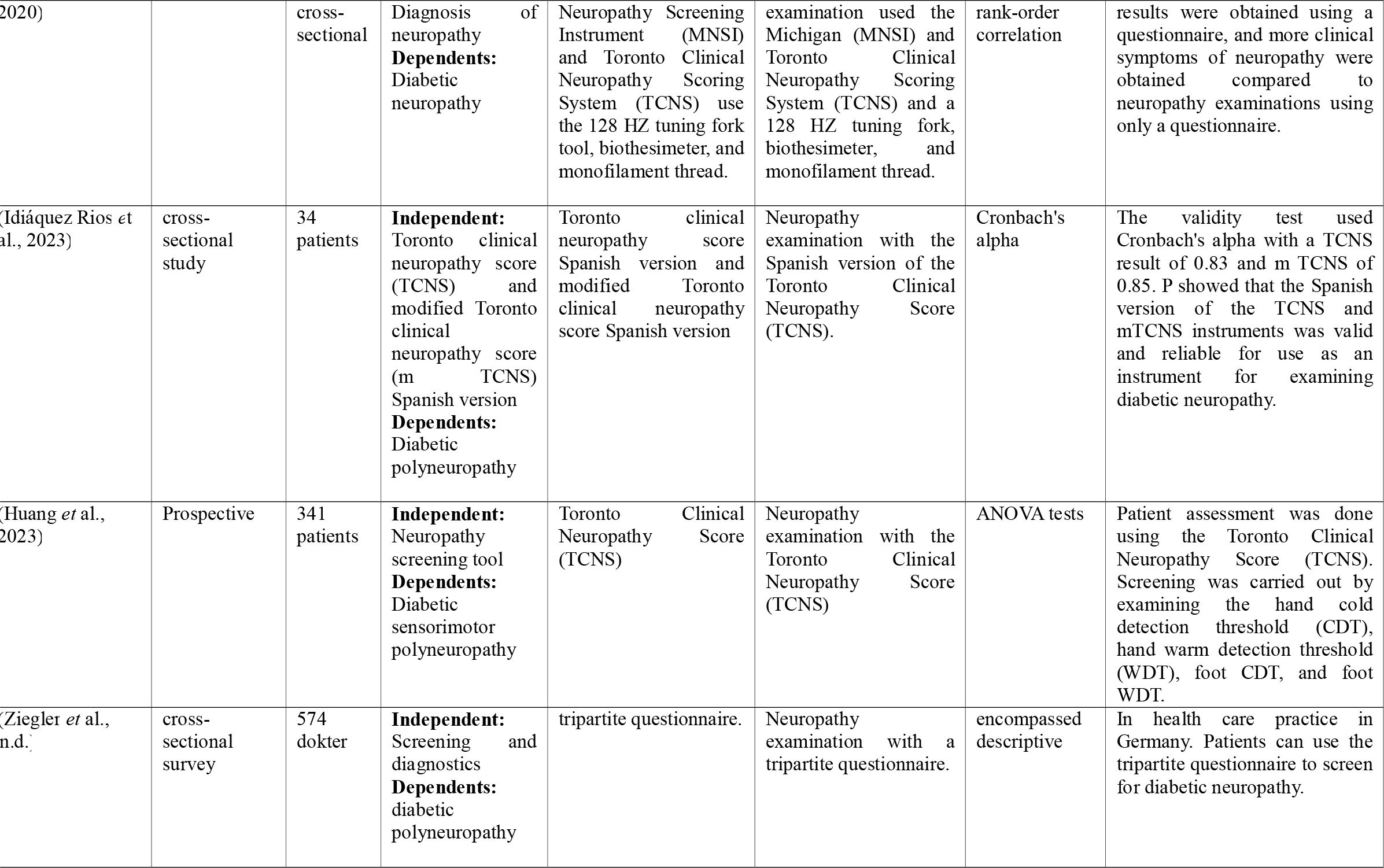
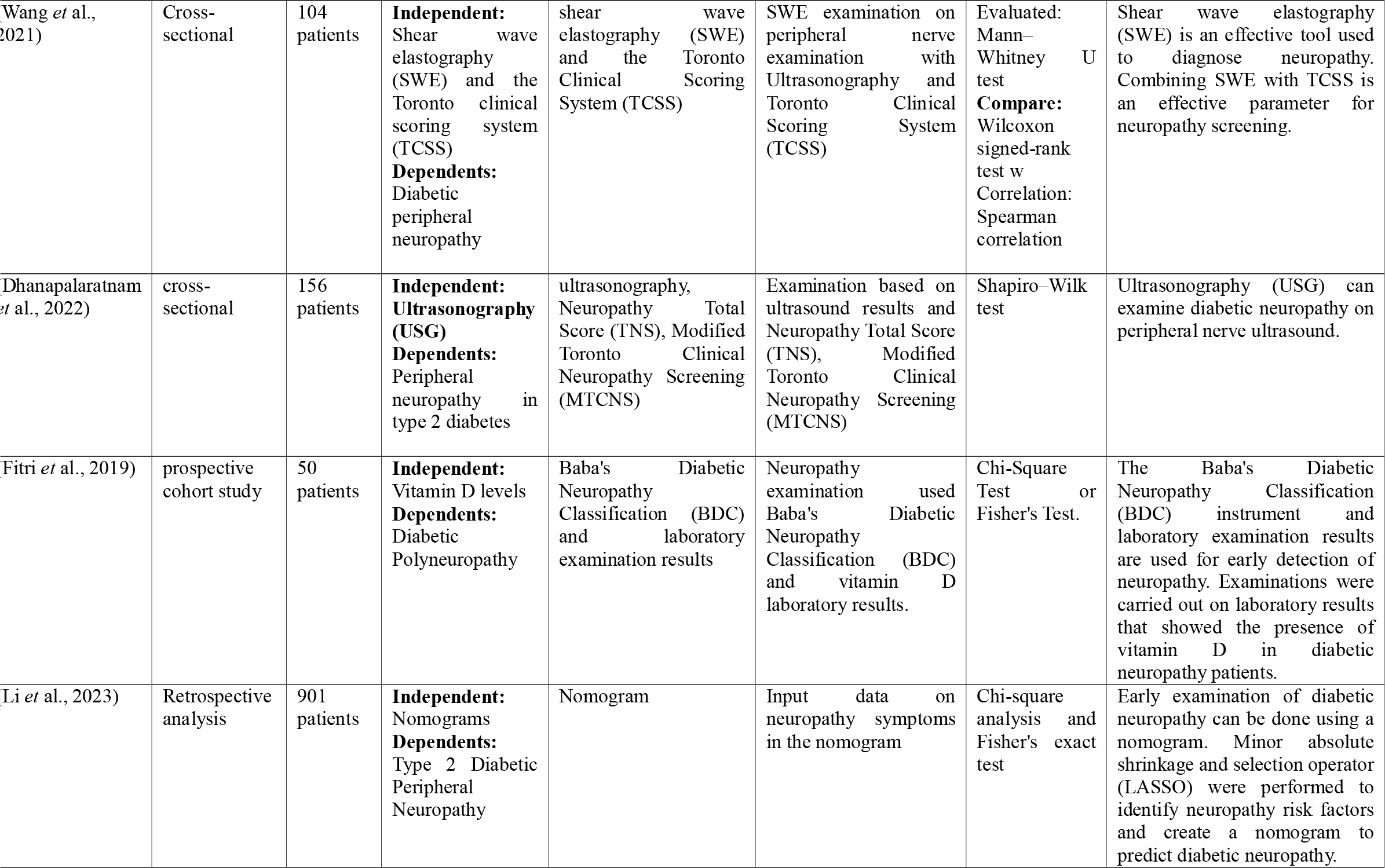
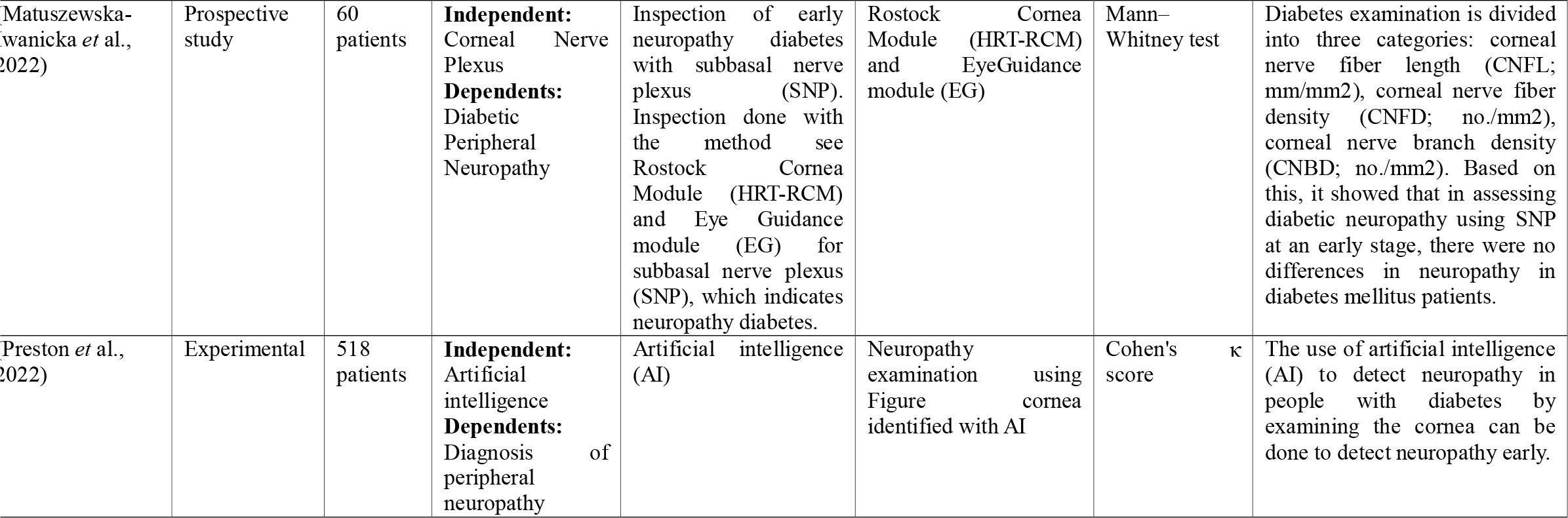
Journal review.

## Discussion

### 1. Early Detection of Diabetic Neuropathy Based on the Health Belief Model

Health workers carry out early detection of neuropathy to make a diagnosis and determine appropriate care. The neuropathy examination uses a neuropathy questionnaire and physical examination tools. Early neuropathy detection questionnaires are the Michigan Neuropathy Screening Instrument (MNSI) and the Toronto Clinical Neuropathy Score (TCNS) (Ramanathan *et* al., 2021). Another study translated the Michigan Neuropathy Screening Instrument (MNSI) questionnaire into Turkish. The Michigan Neuropathy Screening Instrument (MNSI) questionnaire results were divided into three categories: mild neuropathy, moderate neuropathy, and severe neuropathy (Kaymaz *et* al., 2020). Another study used the Toronto Clinical Neuropathy Score (TCNS) questionnaire for early detection of diabetic neuropathy (Huang *et* al., 2023). The Toronto Clinical Neuropathy Score (TCNS) questionnaire detects neuropathy by asking about symptoms, reflex examination, and sensitivity examination (Rios *et* al., 2023).

Nurses carry out early detection of diabetic neuropathy using diabetic neuropathy instruments (Dhanapalaratnam *et* al., 2022). Vibration perception threshold (VPT) examines vibration sensitivity using a neurotesimeter. The examiner applies vibrations to the heel at 200 Hz and the metatarsals at 30 Hz (Drechsel *et* al., 2021). Another article stated that vibrations are given in the stages from small vibrations to more significant vibrations with a sensitivity limit of >14.9 V. Patients who do not feel vibrations >14.9 V showed diabetic neuropathy (Liu *et* al., 2021). Vibration perception threshold (VPT) examination uses a biotesimeter with vibrations of 0-50 V. Neuropathy examination results show adverse neuropathy if the patient feels vibrations <25 V; patients show positive neuropathy if they feel vibrations ≥25 V(Ramanathan *et* al., 2021).

Health workers diagnose diabetic neuropathy using peripheral nerve ultrasonography (USG) in diabetes mellitus patients (Dhanapalaratnam *et* al., 2022). Shear wave elastography (SWE) is an examination of diabetic neuropathy using elastography images (Wang *et* al., 2021). neuropathy examination with Baba’s Diabetic Neuropathy Classification (BDC) questionnaire and vitamin D laboratory results (Fitri *et* al., 2019). Nurses used nomograms to input data on neuropathy symptoms in diabetic patients. Neuropathy symptoms that exceed the nomogram limit indicate diabetic neuropathy (Li *et* al., 2023).

Early neuropathy examination uses Artificial Intelligence (AI) by looking at the corneas of diabetic patients. However, examinations using artificial intelligence (AI) have not been proven to show significant differences in signs of neuropathy in diabetes mellitus patients (Preston *et* al., 2022).

### 2. Ontology Review of Early Detection of Diabetic Neuropathy Based on Health Belief Model for Diabetic Ulcers Prevention

The Health Belief Model was developed by Rosenstock in 1966. The Health Belief Model is used to identify individual behavior to prevent disease (Orbell, 2020). According to Rosenstock, individual behavior toward health is influenced by health concerns, beliefs about disease susceptibility, and beliefs about the consequences of disease (Mckellar & Sillence, 2020). The health belief model identifies behavior to prevent complications in chronic diseases such as diabetes mellitus. Patients apply the health belief model in preventing complications of diabetes mellitus (Wang *et* al., 2023). Diabetic neuropathy is the most frequent complication of diabetes mellitus (Akhtar *et* al., 2023).

The nurse performs a diabetic neuropathy examination to establish a diagnosis and determine appropriate treatment—early detection of neuropathy by examining signs, symptoms, vibration threshold, sensitivity, and reflexes. Instruments for early detection of diabetic neuropathy can be questionnaires and physical examination tools such as monofilaments, biotesimeters, and tuning forks (Liu *et* al., 2021). Early neuropathy diagnosis and proper foot care can prevent diabetic ulcers (Basir *et* al., 2020).

### 3. Epistemology Review of Early Detection of Diabetic Neuropathy Based on Health Belief Model for Prevention of Diabetic Ulcers

The health belief model has five components: perceived susceptibility, perceived severity, perceived benefits, perceived barriers, and cues to action (Kahaleh & Truong, 2021). The behavior of diabetes mellitus patients is based on their beliefs regarding vulnerability, seriousness, obstacles, and perceived benefits. Family support, health services, and availability of instruments influence patient motivation in implementing disease prevention behavior (Wang *et* al., 2023).

Early examination of neuropathy used questionnaires and physical examination. The Michigan Neuropathy Screening Instrument (MNSI) questionnaire consists of 12 questions about neuropathy symptoms (Kaymaz *et* al., 2020). The Toronto Clinical Neuropathy Score (TCNS) questionnaire contains questions about signs of neuropathy symptoms, reflex examination, and sensitivity examination in diabetes mellitus patients (Huang *et* al., 2023). In one article, researchers combined the TCNS questionnaire with other instruments such as shear wave elastography (SWE) (Wang *et* al., 2021). The diagnosis of neuropathy is based on peripheral nerve ultrasound (USG) (Dhanapalaratnam *et* al., 2022).

Vibration perception threshold (VPT) examination using a biotesimeter. The magnitude of the vibrations was 200 Hz on the heel and 30 Hz on the metatarsals. Patients were diagnosed with neuropathy if they did not feel vibrations (Liu *et* al., 2021). Diabetic neuropathy examination used monofilament thread for food sensitivity testing—a nurse examined by inserting a monofilament thread into the patient’s leg. The examination results showed neuropathy if the patient did not feel the prick of the monofilament thread (Oh *et* al., 2022).

Early diabetic neuropathy examination used Baba’s Diabetic Neuropathy Classification (BDC) and vitamin D laboratory results (Fitri *et* al., 2019). Nurses used a nomogram to detect neuropathy by entering data on signs of neuropathy symptoms in the nomogram. Symptom data in the nomogram line showed neuropathy (Li *et* al., 2023)—early examination of neuropathy using artificial intelligence (AI) on images of the cornea of diabetes patients. However, examinations using artificial intelligence (AI) have not been proven to be accurate in diagnosing diabetic neuropathy (Preston *et* al., 2022).

### 4. Review of Axiology Early Detection of Diabetic Neuropathy Based on Health Belief Model for Prevention of Diabetic Ulcers

Early detection of diabetic neuropathy reduces the risk of diabetic ulcers in diabetes mellitus patients. Early detection of neuropathy occurs in 2 ways: examination using a neuropathy questionnaire and physical examination using a neuropathy instrument. The availability of health facilities and patient health beliefs can motivate patients to implement health behavior (Savari *et* al., 2023).

Patients implement behavior to prevent complications by early detection of neuropathy. Nurses diagnose neuropathy early and can determine nursing care for diabetic neuropathy. This behavior can prevent complications of diabetic ulcers (Crowley *et* al., 2023).

## Conclusion

Early detection of diabetic neuropathy is carried out to determine the severity of neuropathy in diabetes mellitus patients. Diabetic neuropathy examination is carried out in 2 ways: a neuropathy questionnaire and a physical examination using a neuropathy instrument. Neuropathy questionnaires are Michigan Neuropathy Screening Instrument (MNSI), Toronto Clinical Neuropathy Score (TCNS), and Diabetic Neuropathy Classification (BDC). Physical examination tools include biotesimeters, vibrations, neurotesimeters, tuning forks, monofilament threads, ultrasonography (USG) results, artificial intelligence (AI), and nomograms.

## Data Availability

All data produced in the present study are available upon reasonable request to the authors

